# Preeclampsia prediction with maternal and paternal polygenic risk scores: the TMM BirThree Cohort Study

**DOI:** 10.1101/2024.02.07.24302476

**Authors:** Hisashi Ohseto, Mami Ishikuro, Taku Obara, Akira Narita, Ippei Takahashi, Genki Shinoda, Aoi Noda, Keiko Murakami, Masatsugu Orui, Noriyuki Iwama, Masahiro Kikuya, Hirohito Metoki, Junichi Sugawara, Gen Tamiya, Shinichi Kuriyama

**Author notes:** **Corresponding author:** Mami ISHIKURO, PhD, 2-1 Seiryo-machi, Aoba-ku, Sendai, Miyagi, 980-8573, Japan.

## Abstract

**Background:** Genomic information from pregnant women and their husbands may provide effective biomarkers for preeclampsia. This study investigated how parental polygenic risk scores (PRSs) for blood pressure (BP) and preeclampsia are associated with preeclampsia onset and evaluated predictive performances of PRSs with clinical predictive variables.

**Methods:** In the Tohoku Medical Megabank Project Birth and Three-Generation Cohort Study, participants were genotyped using either Affymetrix Axiom Japonica Array v2 (further divided into two cohorts—the PRS training cohort and the internal-validation cohort—at a ratio of 1:2) or Japonica Array NEO (external-validation cohort). PRSs were calculated for systolic BP (SBP), diastolic BP (DBP), and preeclampsia. Associations between PRSs and preeclampsia, including preeclampsia superimposed on chronic hypertension, were examined using logistic regression analysis; prediction models were developed using a competing-risks approach with clinical predictive variables and PRSs.

**Results:** In total, 19,836 participants were included. Hyperparameters for PRS calculation were optimized for 3,384 participants in the training cohort. In internal- and external-validation cohorts, 357 of 6,768 (5.3%) and 269 of 9,684 (2.8%) participants developed preeclampsia, respectively. Preeclampsia onset was significantly associated with maternal PRSs for SBP and DBP in internal- and external-validation cohorts and with paternal PRSs for SBP and DBP only in the external-validation cohort. Maternal PRSs for DBP calculated using “LDpred2” most improved prediction models. Maternal PRSs for DBP provided additional predictive information on clinical predictive variables. Paternal PRSs for DBP improved prediction models in the internal-validation cohort.

**Conclusions:** Parental PRS, along with clinical predictive variables, is potentially useful for predicting preeclampsia.

## Introduction

Preeclampsia (PE) is a multisystem disorder characterized by de novo hypertension and proteinuria, affecting approximately 3.4% of pregnant women in the USA^1^ and 2.7% in Japan.^2^ It causes approximately 45,900 maternal deaths worldwide.^3^ There are effective interventions, such as moderate exercise,^4^ aspirin,^5^ and calcium supplementation.^6^ Thus, early detection of high-risk pregnancies can lead to better outcomes^7^ for mothers and fetuses. Therefore, developing accurate PE prediction models is critical in clinical practice.

More than a hundred prediction models for PE are reported,^8^ with some using genomic information from pregnant women (here, “maternal genomic information”).^9–12^ As the heritability of PE was estimated to be 55% in a family study, with maternal and fetal contributions accounting for 35% and 20%, respectively,^13^ maternal genomic information is expected to provide a good biomarker for PE prediction. Integrating maternal genomic information as a polygenic risk score (PRS) can potentially improve existing predictive models; however, relevant research remains insufficient. In previous studies,^9–12^ PRS for blood pressure (BP) and PE were utilized but some important clinical predictive variables such as actual measured BP values and family history of PE were not simultaneously incorporated into the prediction models. Actual measured BP values in early pregnancy is an effective PE biomarker and is included in many clinical prediction models.^8, 14^ Similarly, family medical history is a predictive variable for diseases and reflects the genetic load and shared environmental factors.^15, 16^ A recent study revealed that the PRS and family medical history provide complementary information on noncommunicable diseases.^17^ As actual measured BP values and family history of hypertensive disorders of pregnancy (HDP) are more accessible than PRS, their combined clinical usefulness with genomic information should be explored. A previous study^18^ reported no improvement when PRS was incorporated into a prediction model with clinical predictive variables using machine learning. However, this study had a small number of cases (< 100) and was not externally validated.

Given that genomic information from pregnant women’s husbands (here, “paternal genomic information”) is transmitted to both fetal and placental tissues, paternal genomic information holds predictive potential for PE. As obtaining fetal genome is difficult, combining the maternal and paternal genomes may capture a comprehensive genetic predisposition to PE. However, no previous studies have employed such information in PE prediction models.

Here, we examined the relationship between genomic information from pregnant women and their husbands (here, “parental genomic information”), and PE onset. Moreover, we investigated whether parental PRSs have predictive information in addition to clinical models, including actual measured BP values and family history of HDP.

## Materials and Methods

### Participants

The Tohoku Medical Megabank Project Birth and Three-Generation Cohort Study (the BirThree Cohort Study)^19, 20^ recruited pregnant women and their families between 2013 and 2017. More than 50 obstetric clinics and hospitals in the Miyagi Prefecture, Japan participated, registering 23,406 pregnant women and their 8,823 husbands.

We excluded the following pregnant women from the study: those who withdrew consent, had multiple pregnancies, had stillbirth before 20 weeks of gestation, and missed the diagnosis of PE or delivery date. Here, we included only the first-time participations of those who had multiple participations in the TMM BirThree Cohort Study.

Participants were genotyped using either the Affymetrix Axiom Japonica Array v2 (JPA v2) or Japonica Array NEO (JPA NEO). Those with missing genotyping data were excluded after a standard quality control procedure.^21^ Those genotyped using JPA v2 were divided into two cohorts—the PRS training cohort and the maternal internal validation cohort—at a ratio of 1:2. Those genotyped using JPA NEO were defined as the maternal external validation cohort. The participants’ husbands were genotyped using the same genotyping array platform as the participants. Cohorts with participants’ husbands’ genotypes in the maternal internal and external validation cohorts were defined as the parental internal and external validation cohorts, respectively (Figure 1). Ethical approval was obtained from the Ethics Committee of the Tohoku Medical Megabank Organization (2023-4-025), and informed consent for research participation was obtained from all participants.

**FIGURE 1.**
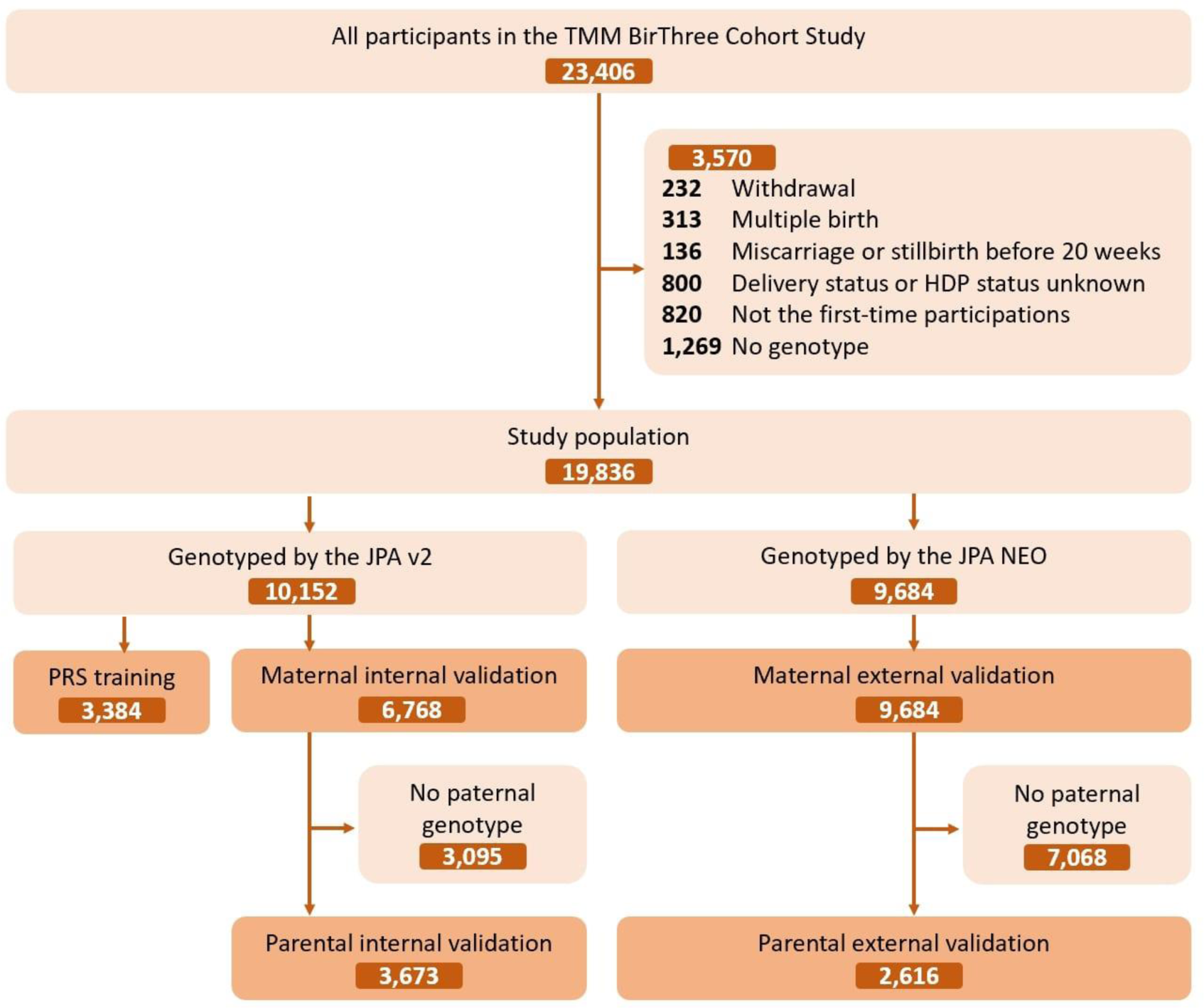
TMM BirThree Cohort Study flow for three study cohorts. After excluding ineligible participants, the cohort was divided into three cohorts, namely PRS training, maternal internal validation, and maternal external validation. The cohorts with participants’ husbands’ genotypes in the maternal internal and external validation cohorts were the parental internal and external validation cohorts, respectively. TMM BirThree Cohort Study: The Tohoku Medical Megabank Project Birth and Three-Generation Cohort Study. JPA v2: Affymetrix Axiom Japonica Array v2; JPA NEO: Affymetrix Axiom Japonica Array NEO.

### Predictive variables

Based on previous research on PE prediction models,^22–26^ we selected following predictive variables: maternal age at conception, pre-pregnancy body mass index (BMI), chronic hypertension (CH), systemic lupus erythematosus (SLE), type 1 and 2 diabetes mellitus (DM), maternal family (mother or sisters) history of HDP, conception via in vitro fertilization (IVF), parity (nulliparous, parous with or without previous HDP), gestational age at the previous delivery, the inter-birth interval, and BP at the first antenatal care during 10–13 weeks of gestation. We collected BP data at 10–13 weeks of gestation, when the majority of population underwent the first or second antenatal care. Mean arterial pressure (MAP) was calculated and converted to log_10_ transformed multiple of the median (log MoM) for the prediction model. Paternal age and family history of HDP were obtained for paternal analyses.

### Polygenic risk score

Genotyping and PRS calculations are described in Supplementary Methods. In brief, PRSs for three phenotypes, systolic BP (SBP), diastolic BP (DBP), and PE, were calculated using two methods, genome-wide clumping and thresholding (C+T) and a Bayesian approach using LDpred2.^27, 28^ The hyperparameters were optimized in the PRS training cohort.

### Outcome measurement

PE was identified based on the American College of Obstetricians and Gynecologists (ACOG) guidelines from 2002^29^ using medical records at antenatal care. PE superimposed on the CH was included in the definition of PE. PE was automatically diagnosed using an algorithm based on medical record data and validated by a physician. The details are given elsewhere.^23^

### Model development

We applied the competing risk model,^30^ which has been validated internally and externally in Europe^31^ and yielded comparable results in the TMM BirThree Cohort Study.^22, 23^ The competing risks model assumes that all pregnant women will develop PE during pregnancy, but only some will actually develop PE because of competing of delivery without PE. We employed a parametric survival model with a Gaussian distribution.^30^ Delivery without PE was considered as censored. Missing predictive variables were imputed using k-nearest neighbor imputation with k = 140 (square root of the total study population^32^). To assess the model discrimination, Harrell’s C-statistic was calculated for whole gestational age. To assess model calibration, the calibration slope was calculated by regressing observed survival outcomes on predicted gestational age at delivery with PE in the same parametric survival models.^33^

### Statistical analysis

Baseline characteristics and PE status were compared between cohorts. *P*-values were calculated using the *t*-test for continuous variables and chi-square test or Fisher’s exact test for categorical variables. Pearson’s correlation coefficients were calculated for all combinations of parental PRSs using LDpred2 in the parental internal validation cohort.

Association analyses were performed using SBP-PRS, DBP-PRS, and PE-PRS using LDpred2. In the maternal internal and external validation cohorts, the associations between maternal PRSs and PE onset were examined using logistic regression analysis adjusted for maternal age and four genetic principal components (PC). Two models were developed, one with PRSs as continuous values and the other as tertile values. In the parental internal and external validation cohorts, associations between both maternal and paternal PRSs and PE onset were examined using logistic regression analysis adjusted for parental age and four genetic PCs. The results of internal and external validation cohorts were merged using inverse-variance weighting for meta-analysis. Interactions between maternal and paternal PRSs were also examined. As subanalyses, participants with CH were excluded from the study population, and analyses were performed again.

The prediction model was developed in two stages: identification of a suitable PRS for prediction models and development of clinical prediction models using parental PRSs. In the first stage, prediction models were developed using each maternal PRS, age, and four genetic PCs. Based on the C-statistics of prediction models, we selected either C+T or LDpred2 as the PRS calculation algorithm, and SBP-PRS, DBP-PRS, or PE-PRS as the PRS phenotype. In the second stage, we created four models: reference model, family-history model, at-pregnancy-confirmation model, and in-early-pregnancy model (Supplementary Method). C-statistics and calibration slopes were calculated for each model. The 95% confidence intervals (CI) and internal validation used the bootstrap method with 1,000 resamplings.

All analyses were conducted using R (4.1.0) unless otherwise noted. We considered a two-tailed *P*-value <0.05 as significant.

## Results

A total of 19,836 participants were eligible for the present study: 3,384 in the PRS training cohort, 6,768 in the maternal internal validation cohort, and 9,684 in the maternal external validation cohort (Figure 1). In the maternal internal and external validation cohorts, 3,673 (54.3%) and 2,616 (27.0%) participants, respectively, had paternal genotyping data.

In total, 357 of 6,768 (5.3%) participants and 269 of 9,684 (2.8%) participants developed PE in the maternal internal and external validation cohorts, respectively (Table 1). Of the 3,673 and 2,616 participants in the parental internal and external validation cohorts, respectively, 221 (6.0%) and 23 (0.9%) participants developed PE (Supplementary Table 1). Participants with PE tended to have an earlier delivery; were older; had a higher BMI; had a history of CH, DM, and SLE; were nulliparous or parous with previous PE; had a shorter last delivery gestational age; conceived via IVF; and had a higher BP at 10–13 weeks of gestation, with slight variations among the maternal and parental cohorts (Tables 1 and Supplementary Table 1). Notable differences in the characteristics between the internal and external validation cohorts were observed (Supplementary Tables 2 and 3). Strong correlations existed between maternal PRSs or among paternal PRSs, whereas weak correlations existed between maternal and paternal PRSs (Supplementary Figure 1).

**TABLE 1.**
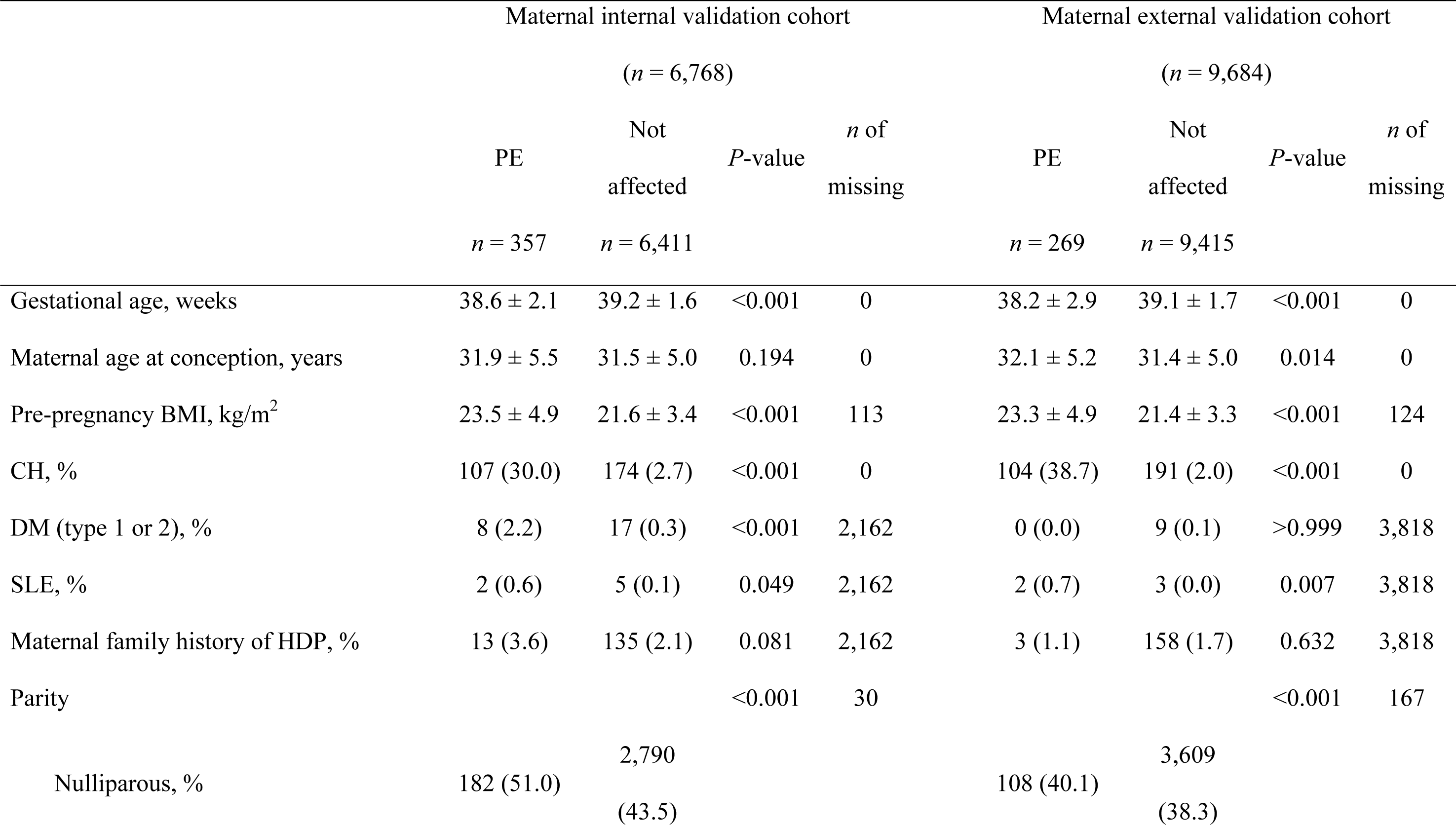
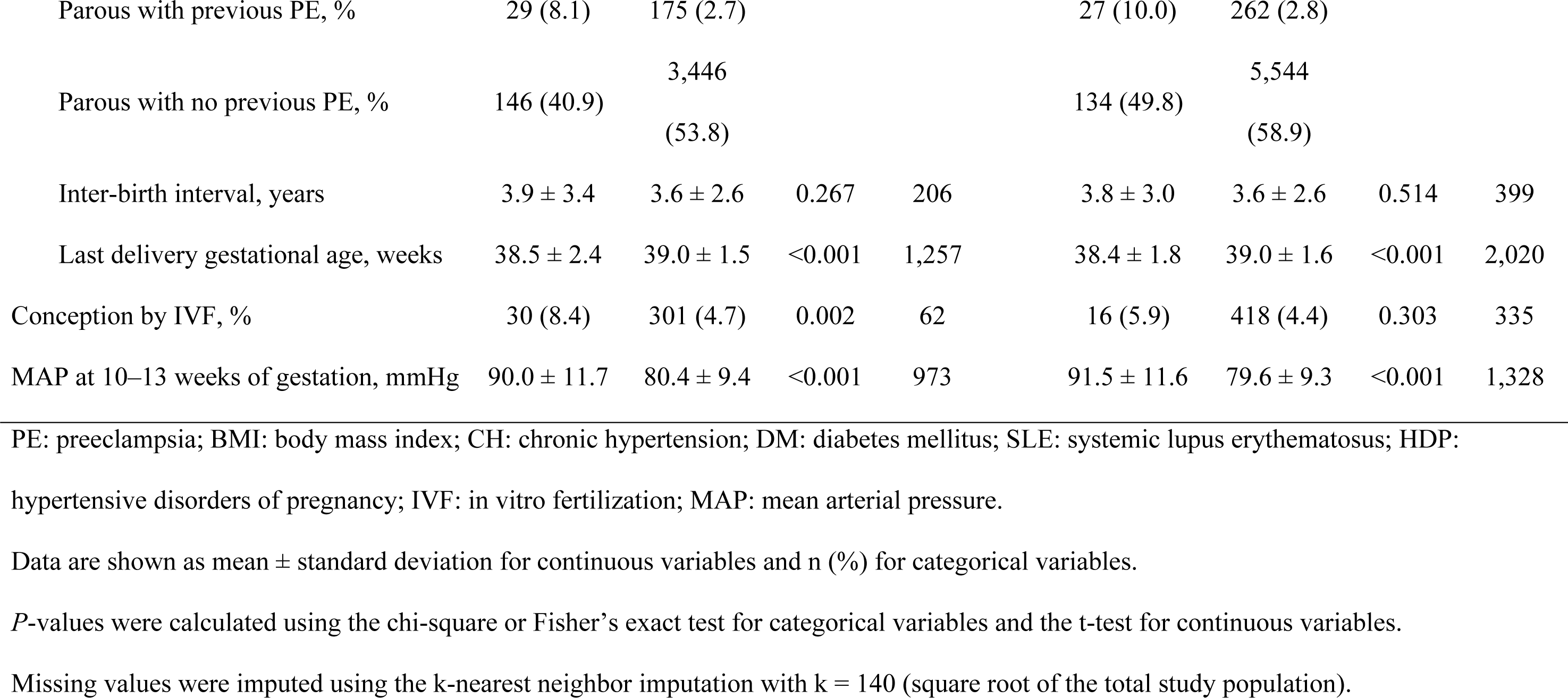
Baseline characteristics in the maternal internal and external cohort, stratified by the PE status.

In association studies for the maternal cohorts (Table 2), maternal SBP-PRS and DBP-PRS were significantly associated with PE onset in the meta-analysis; odds ratio (OR) and 95% CI per 1 standard deviation (SD): 1.17 (1.08–1.27) and 1.21 (1.11–1.31), respectively. In association studies for the parental cohorts (Supplementary Table 4), maternal and paternal PRSs were not related to PE onset in meta-analysis. Only in parental external validation cohort, maternal DBP-PRS, paternal SBP-PRS, and paternal DBP-PRS were associated with PE onset; OR and 95% CI per 1 SD: 1.79 (1.14–2.81), 1.97 (1.28–3.04), and 2.04 (1.32–3.17), respectively. Similar results were observed after excluding participants with CH. However, the results did not converge in the parental external validation cohort because of the small number of outcomes (Supplementary Tables 5 and 6). No interaction was noted between the maternal and paternal PRSs (data not shown).

**TABLE 2.** Relationship between maternal PRSs and PE onset in the maternal cohorts by logistic regression analysis.

|  | Internal validation cohort |  | External validation cohort |  | Meta-analysis |  |
| --- | --- | --- | --- | --- | --- | --- |
|  | OR (95% CI) | *P*-value | OR (95% CI) | *P*-value | OR (95% CI) | *P*-value |
| PRS for SBP |  |  |  |  |  |  |
| Continuous | 1.05 (0.94–1.17) | 0.399 | 1.36 (1.20–1.54) | <0.001 | 1.17 (1.08–1.27) | <0.001 |
| Tertile 1 | Reference |  | Reference |  | Reference |  |
| Tertile 2 | 1.09 (0.83–1.45) | 0.525 | 1.06 (0.78–1.44) | 0.712 | 1.08 (0.88–1.33) | 0.472 |
| Tertile 3 | 1.02 (0.78–1.34) | 0.867 | 1.74 (1.30–2.32) | <0.001 | 1.31 (1.07–1.60) | 0.008 |
| PRS for DBP |  |  |  |  |  |  |
| Continuous | 1.09 (0.98–1.22) | 0.119 | 1.38 (1.22–1.57) | <0.001 | 1.21 (1.11–1.31) | <0.001 |
| Tertile 1 | Reference |  | Reference |  | Reference |  |
| Tertile 2 | 0.77 (0.57–1.03) | 0.082 | 1.39 (1.02–1.88) | 0.036 | 1.02 (0.83–1.26) | 0.835 |
| Tertile 3 | 1.09 (0.84–1.42) | 0.498 | 2.20 (1.63–2.96) | <0.001 | 1.48 (1.22–1.80) | <0.001 |
| PRS for PE |  |  |  |  |  |  |
| Continuous | 0.89 (0.80–1.00) | 0.047 | 1.14 (1.01–1.28) | 0.030 | 1.00 (0.92–1.09) | 0.961 |
| Tertile 1 | Reference |  | Reference |  | Reference |  |
| Tertile 2 | 0.94 (0.73–1.21) | 0.625 | 1.45 (1.07–1.95) | 0.016 | 1.12 (0.92–1.36) | 0.241 |
| Tertile 3 | 0.80 (0.62–1.05) | 0.111 | 1.21 (0.89–1.64) | 0.231 | 0.96 (0.78–1.17) | 0.677 |

PRS: polygenic risk score; PE: preeclampsia; OR: odds ratio; CI: confidence interval; SBP: systolic blood pressure; DBP: diastolic blood pressure.
Two models were developed, one with PRS as a continuous value and the other as tertile values.
All results were adjusted for maternal age at conception and four genetic principal components.

After parameter optimization in the PRS training cohort, the predictive performances of SBP-, DBP-, and PE-PRS calculated using the two methods, C+T and LDpred2, were compared in the maternal internal and external validation cohorts (Table 3). The DBP-PRS calculated using LDpred2 improved the discrimination performance the most in both the internal (0.568 in the model with PRS vs. 0.562 in the model without PRS) and external (0.602 in the model with PRS vs. 0.555 in the model without PRS) validation.

**TABLE 3.**
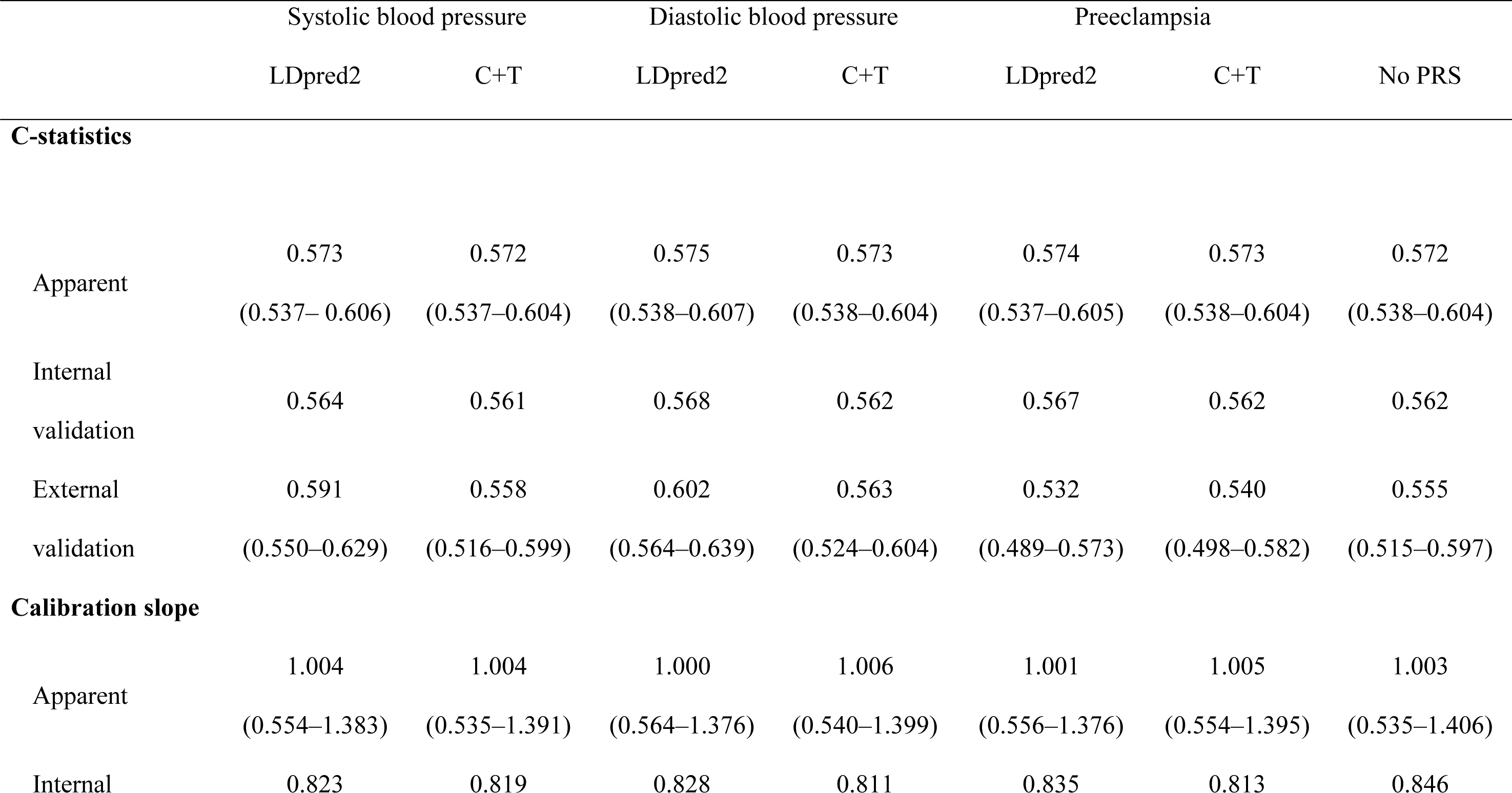
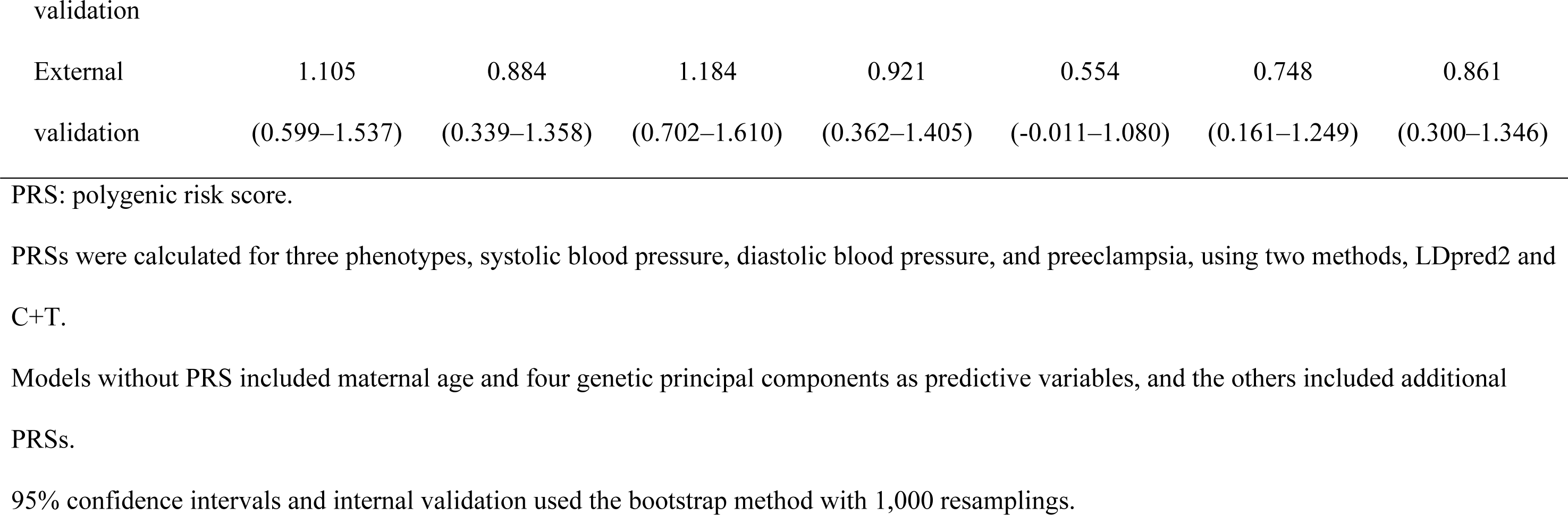
Prediction performance with maternal PRS derived from three phenotypes using two methods.

|  | Systolic blood pressure |  | Diastolic blood pressure |  | Preeclampsia |  |  |
| --- | --- | --- | --- | --- | --- | --- | --- |
|  | LDpred2 | C+T | LDpred2 | C+T | LDpred2 | C+T | No PRS |
| **C-statistics** |  |  |  |  |  |  |  |
| Apparent | 0.573 | 0.572 | 0.575 | 0.573 | 0.574 | 0.573 | 0.572 |
|  | (0.537–0.606) | (0.537–0.604) | (0.538–0.607) | (0.538–0.604) | (0.537–0.605) | (0.538–0.604) | (0.538–0.604) |
| Internal validation | 0.564 | 0.561 | 0.568 | 0.562 | 0.567 | 0.562 | 0.562 |
| External validation | 0.591 | 0.558 | 0.602 | 0.563 | 0.532 | 0.540 | 0.555 |
|  | (0.550–0.629) | (0.516–0.599) | (0.564–0.639) | (0.524–0.604) | (0.489–0.573) | (0.498–0.582) | (0.515–0.597) |
| **Calibration slope** |  |  |  |  |  |  |  |
| Apparent | 1.004 | 1.004 | 1.000 | 1.006 | 1.001 | 1.005 | 1.003 |
|  | (0.554–1.383) | (0.535–1.391) | (0.564–1.376) | (0.540–1.399) | (0.556–1.376) | (0.554–1.395) | (0.535–1.406) |
| Internal | 0.823 | 0.819 | 0.828 | 0.811 | 0.835 | 0.813 | 0.846 |
| External | 1.105 | 0.884 | 1.184 | 0.921 | 0.554 | 0.748 | 0.861 |
| validation | (0.599–1.537) | (0.339–1.358) | (0.702–1.610) | (0.362–1.405) | (-0.011–1.080) | (0.161–1.249) | (0.300–1.346) |

PRS: polygenic risk score.
PRSs were calculated for three phenotypes, systolic blood pressure, diastolic blood pressure, and preeclampsia, using two methods, LDpred2 and C+T.
Models without PRS included maternal age and four genetic principal components as predictive variables, and the others included additional PRSs.
95% confidence intervals and internal validation used the bootstrap method with 1,000 resamplings.

Thereafter, clinical prediction models were developed with maternal and paternal DBP-PRSs calculated using LDpred2. In the maternal cohort, models including maternal PRS demonstrated superior predictive discrimination compared to models without maternal PRS in the reference and family-history models in internal validation. This was also observed in all four external validation models, namely the reference, family-history, at-pregnancy-confirmation, and in-early-pregnancy models (Table 4). For example, in external validation, the C-statistics of the at-pregnancy-confirmation model were 0.778 without PRS and 0.788 with maternal PRS, indicating the utility of maternal PRS at pregnancy confirmation. Even in the in-early-pregnancy model, which included BP in early pregnancy, the C-statistics improved slightly by including maternal PRS (0.841 without PRS vs. 0.844 with maternal PRS), suggesting the utility of maternal PRS in the prediction of early pregnancy. In the parental internal validation cohort (Table 5), paternal PRS, but not maternal PRS, improved the reference, family-history, and at-pregnancy-confirmation models. In the parental external validation cohort, maternal, but not paternal, PRS improved the reference and family-history models, although C-statistics and calibration had a wide 95% CI and the results were not stable. Models with both maternal and paternal PRSs did not demonstrate better discrimination ability than those with either maternal or paternal PRS (Table 5).

**TABLE 4.**
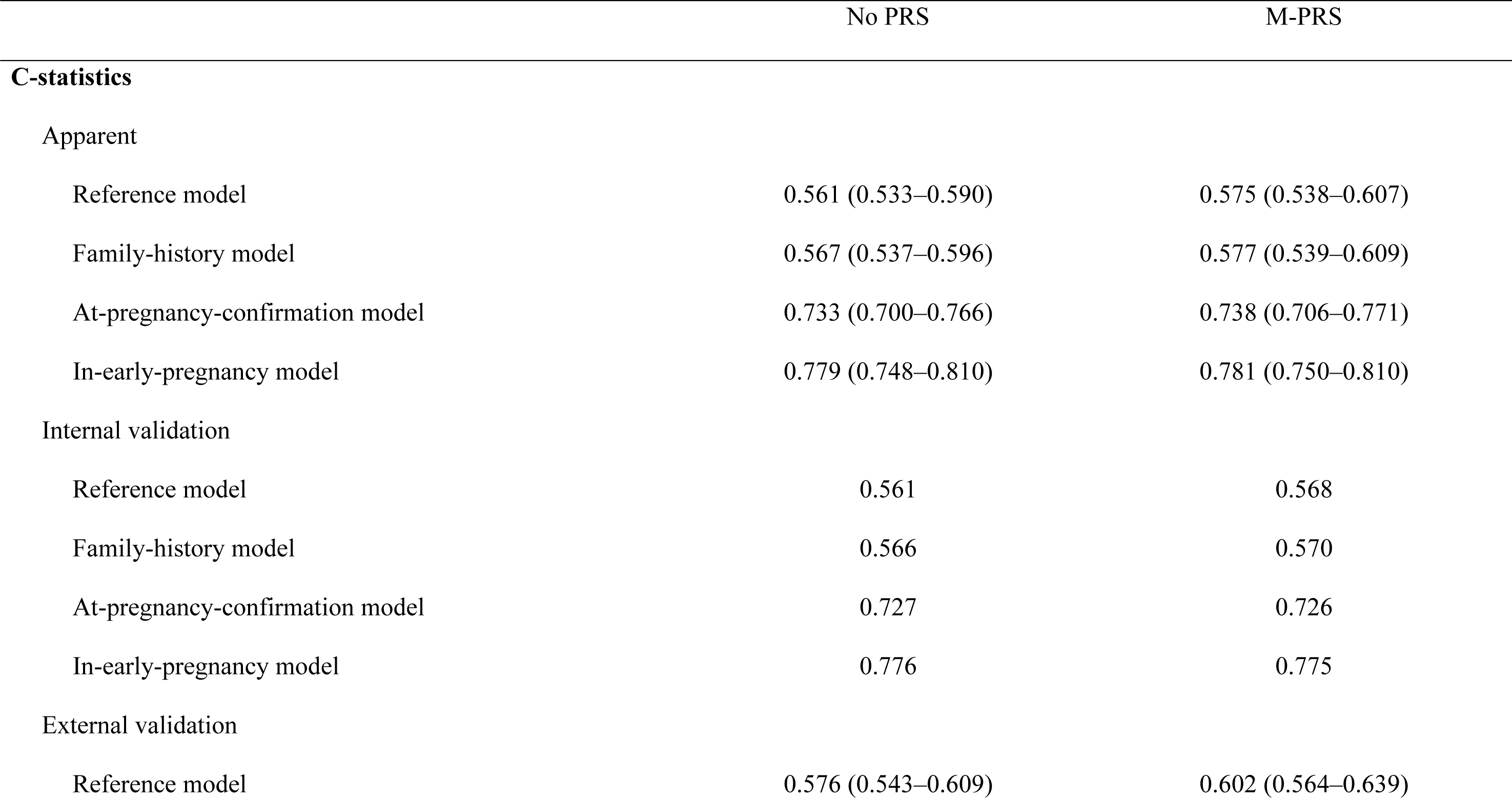
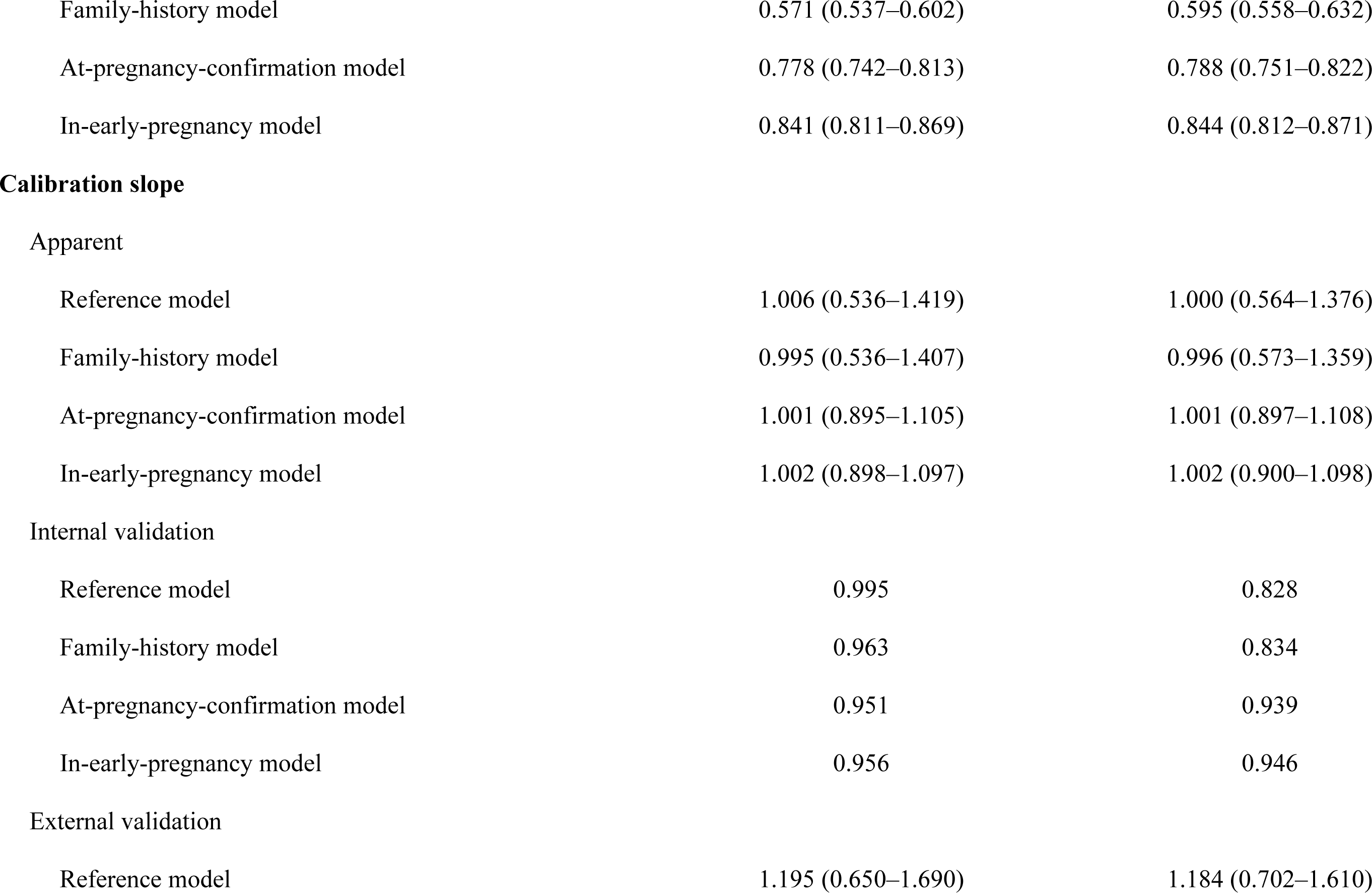
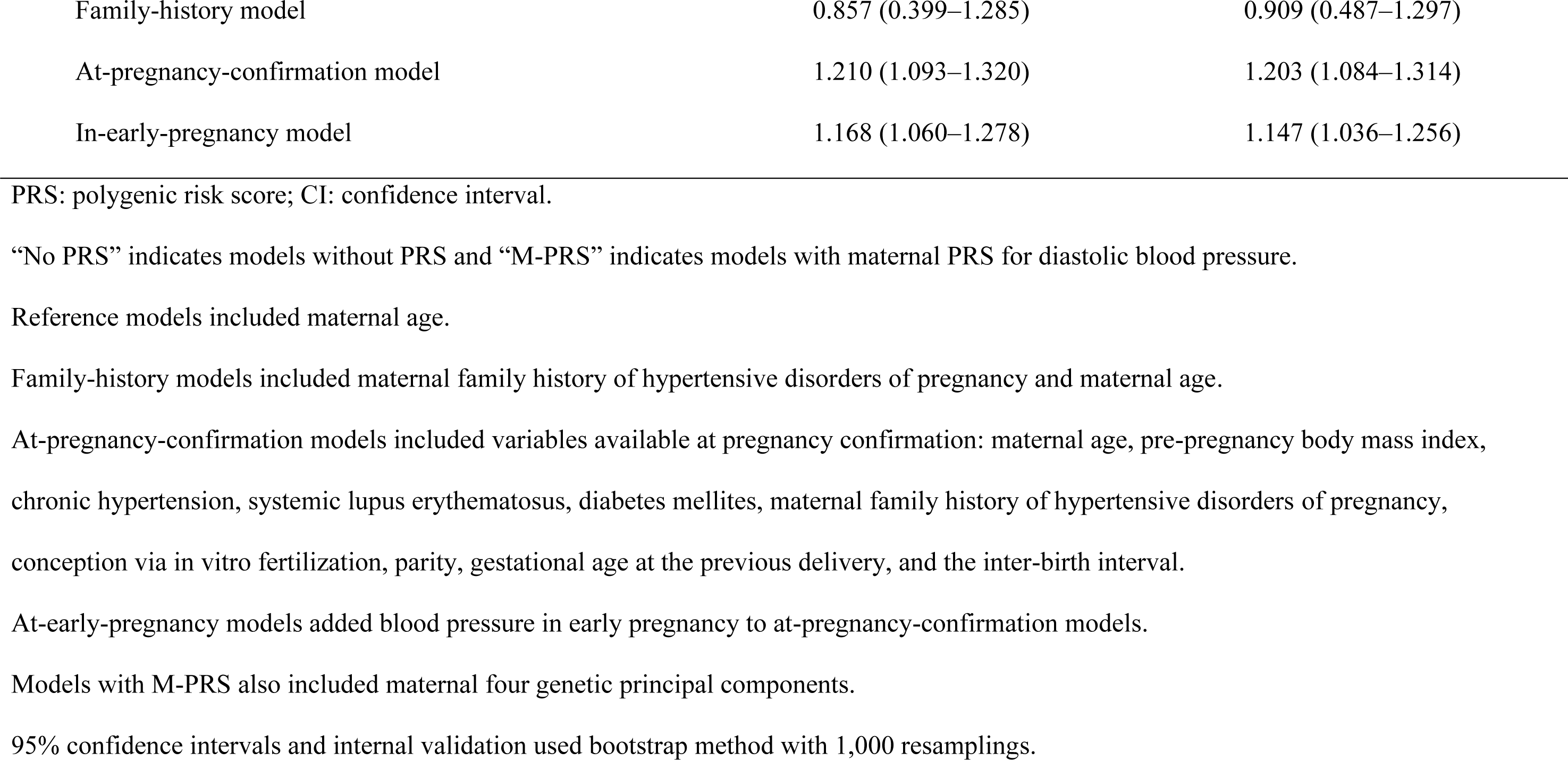
Performance of prediction models with maternal PRS, baseline characteristics, and blood pressure in early pregnancy in the maternal.

**TABLE 5.**
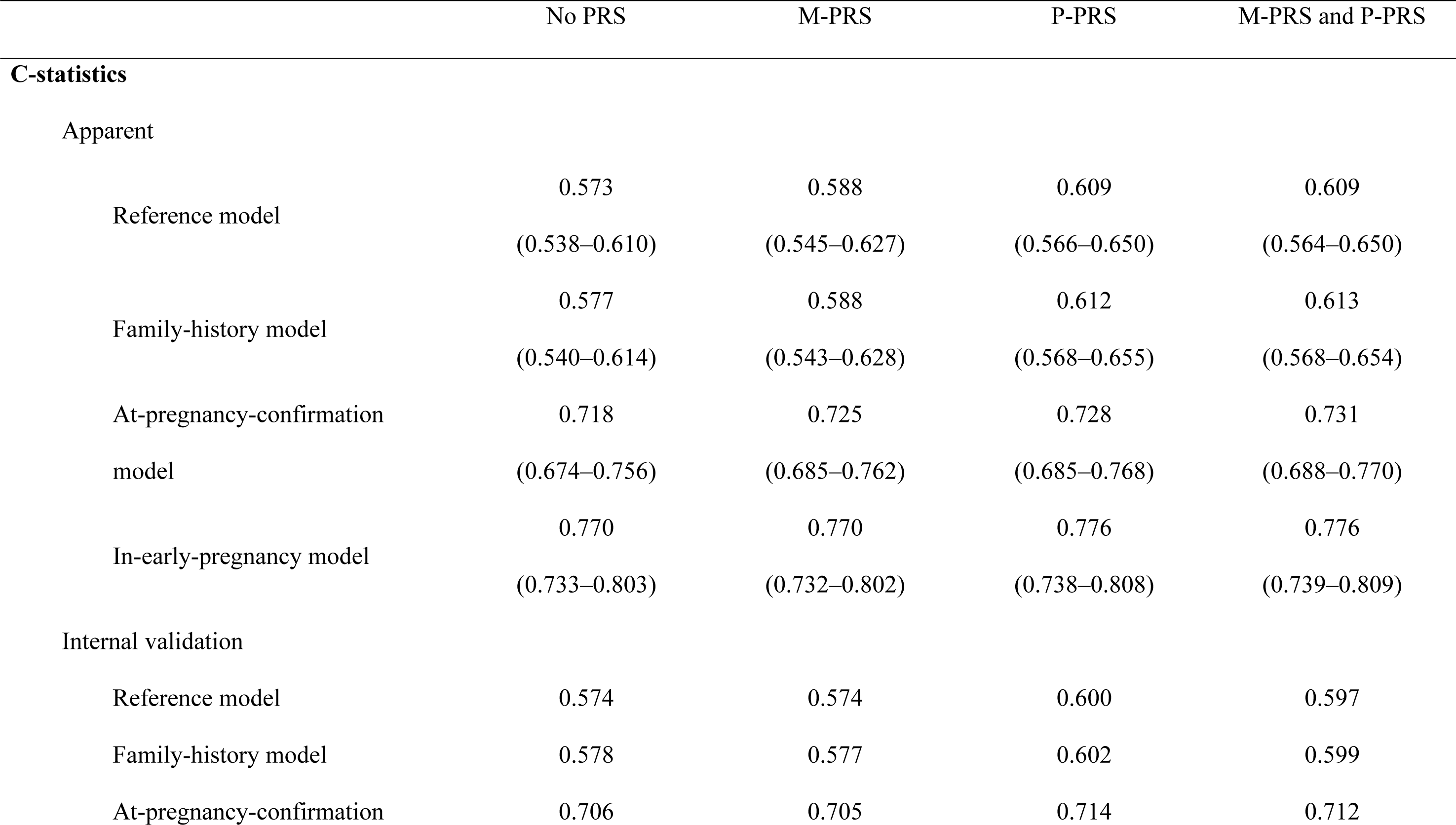
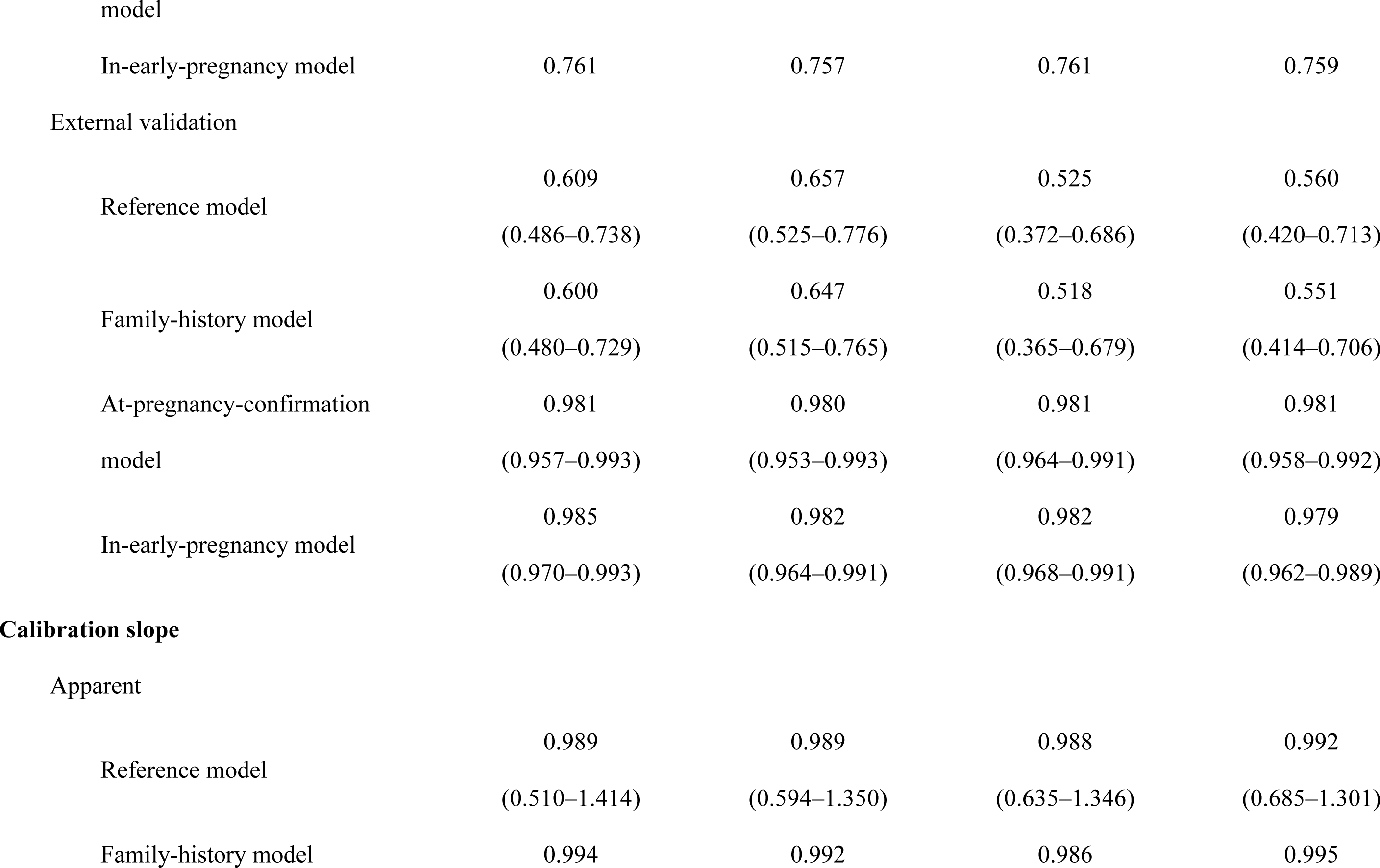
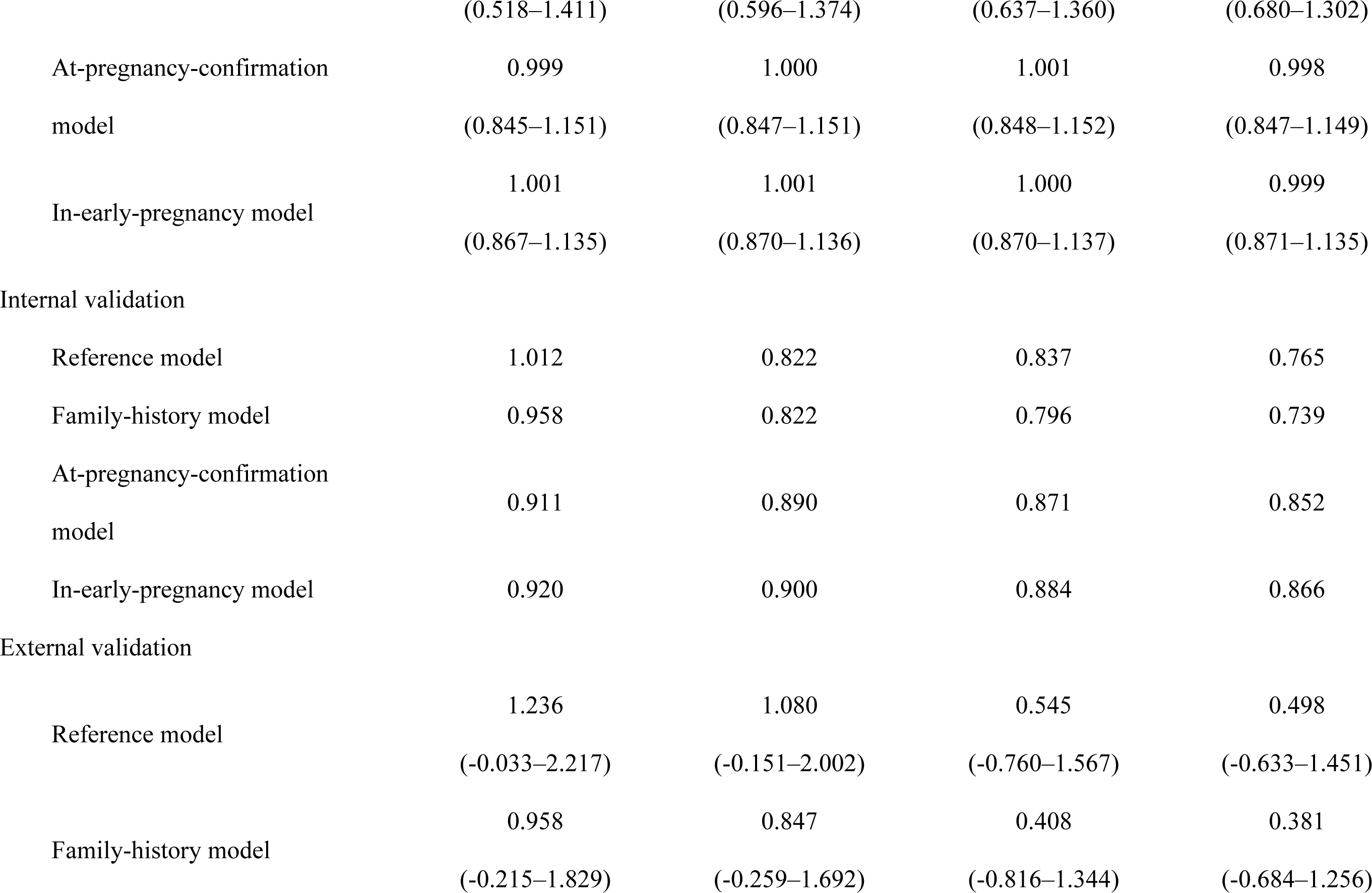
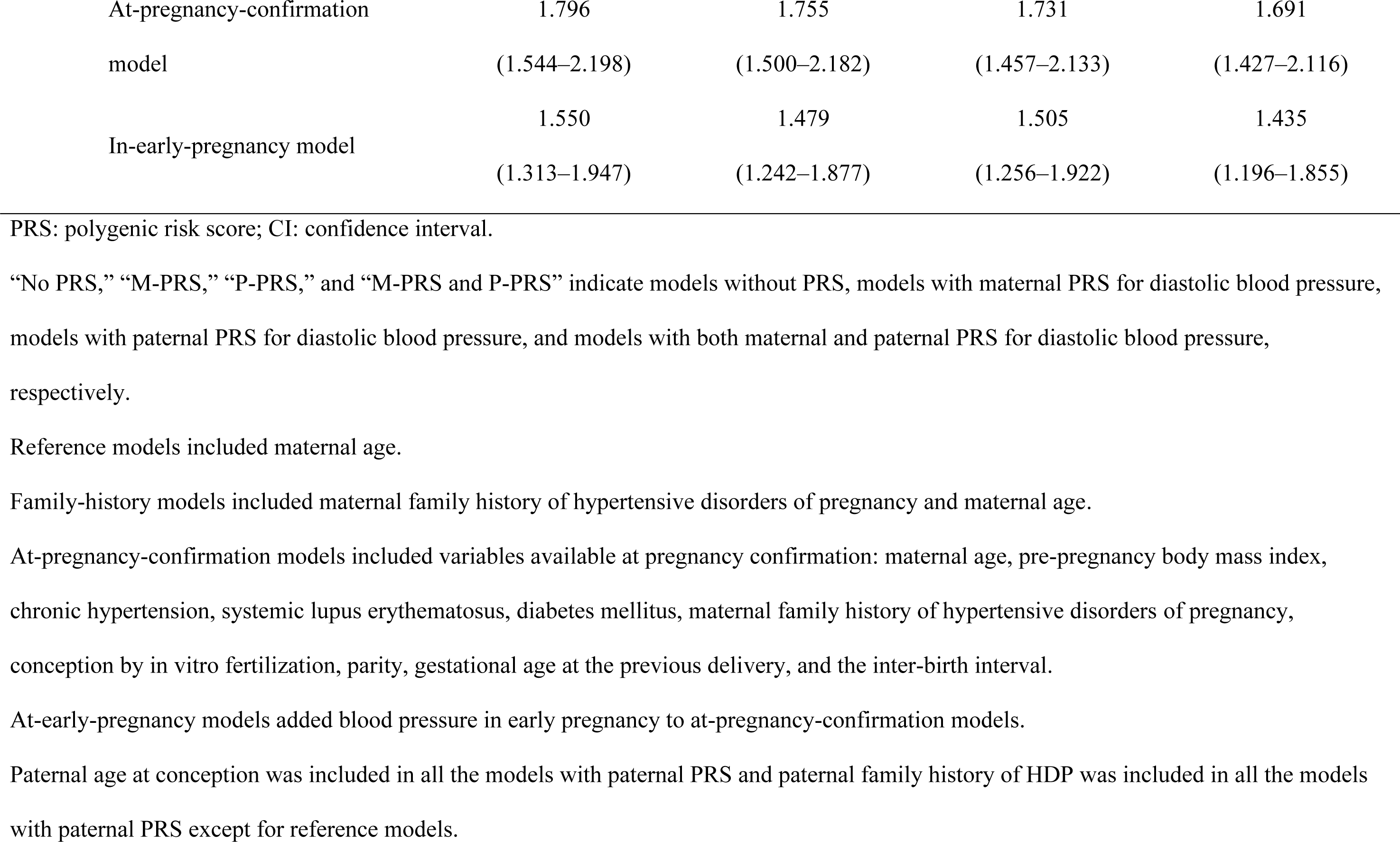
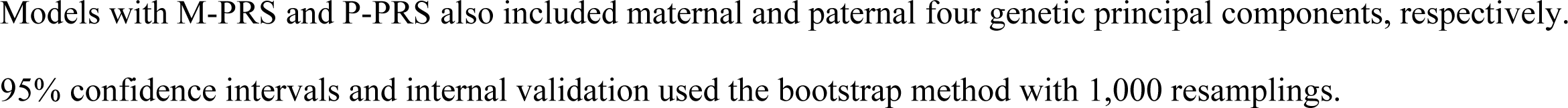
Prediction performance with parental PRS, baseline characteristics, and blood pressure in early pregnancy in the parental cohorts.

## Discussion

The maternal SBP- and DBP-PRSs were consistently associated with PE onset in the internal and external validation cohorts. The paternal SBP- and DBP-PRSs were associated with PE onset only in the external validation cohort. Maternal DBP-PRS calculated using LDpred2 improved prediction models the most. Maternal DBP-PRS improved both at-pregnancy-confirmation and in-early-pregnancy models, indicating the clinical utility of maternal PRS. Paternal PRS improved prediction models in the parental internal validation cohort.

The present study shows that maternal PRSs for BP were associated with PE onset in both internal and external validation cohorts, consistent with previous studies in the European population,^9, 10^ indicating cross-ethnicity generalizability. Moreover, maternal PRS improved the prediction model beyond a family history of HDP. While family history provides plentiful information on genetic predisposition and environment,^15^ the amount of information is determined by family structure and relationships, as well as by disease prevalence. Owing to the demographic trend toward smaller families^34^ and the need to obtain data retrospectively because of the transient nature of the condition during pregnancy, obtaining a comprehensive family history of HDP is difficult. Less than 2% of the participants and less than 1% of participants’ husbands reported a family history of HDP in our cohort. Moreover, reliance only on binary categorization of family history may result in overlooking of the intricate genetic predisposition. Therefore, incorporating PRSs into risk prediction models in addition to family history is crucial for precision medicine. Maternal PRS improved at-pregnancy-confirmation model and slightly improved the in-early-pregnancy model. This indicates that the maternal PRS is useful in clinical practice at pregnancy confirmation and even in early pregnancy, although its predictive significance is diminished by the fact that some genetic predisposition for high BP may manifest itself as the actual measured BP values in early pregnancy. In contrast, PE-RPS was not associated with the development of PE in this meta-analysis. The GWAS for PE^35^ used in our study had a relatively small number of cases, and the number of East Asian populations in the GWAS was limited (123 in the BioBank Japan, 1031 in the UK Biobank, and 1,324 in the FinnGen), which might result in failure to reflect genetic PE predisposition.

Furthermore, in this study, paternal PRSs for BP were associated with PE onset in the external validation cohort. The paternal genetic contribution to PE onset has been predicted in family^13^ and genetic studies.^36^ However, as of date, the association between paternal PRS and PE onset has not been investigated. Our results were inconsistent between the internal and external validation cohorts. One possible reason for this is the biological differences between the parental internal and external validation cohorts. Placental and maternal factors are involved in the development of PE.^37^ The baseline characteristics of the parental internal and external cohorts were quite different; the prevalence of CH in participants with PE was much higher in the external validation cohort than in the internal validation cohort (91.3% vs. 24.9%). This may have led to differences between the two cohorts in terms of the ratio of placental to maternal contributions, resulting in different associations between paternal PRSs and PE. Paternal PRS improved the prediction models in the parental internal validation cohort, despite no significant association in the association analyses. The prediction models developed in the parental internal validation cohort did not have predictive power in the parental external validation cohort possibly, because of cohort differences, as mentioned previously. This suggests that the contribution of paternal PRS to PE onset may differ across populations and should be considered when developing predictive models.

### Strengths and Limitations

Our study is the first to investigate the utility of maternal and paternal PRS for PE prediction. We applied a well-validated prediction model; therefore, our results are close to clinical application. Two cohorts with genomic data from different genotype array platforms and baseline characteristics enabled us to examine the generalizability of the results.

Despite these strengths, our study had four limitations. First, our model did not include effective biomarkers specific to PE, such as the uterine artery pulsatility index and placental growth factor,^25^ which may have improved the performance. However, given the widespread use of genomic information beyond perinatal diseases, prediction models using PRS may be more clinically applicable than those using disease-specific biomarkers. Second, there were several differences between the internal and external validation cohorts. The external validation cohort included more participants with CH, which was the strongest predictive variable, than the internal validation cohort, resulting in calibration slopes >1 in the external validation. Moreover, in the external cohort, a family history of HDP was not associated with PE, limiting the purpose of this study to confirm the predictive value of the PRS in addition to a family history of HDP. Third, because there is no paternal genome-wide association study (GWAS) on PE, we had no choice but to apply the maternal GWAS to the fathers. Fourth, owing to the small number of parental cohorts, the effectiveness of paternal PRS in predicting PE may have been overlooked. Optimization was conducted only for maternal PRS, which may have resulted in an underestimation of the ability of paternal PRS to predict PE.

### Perspectives

Genomic information remains unchanged and can be used throughout life. This is a crucial difference from other biomarkers,^25^ which are only reflective of temporary conditions. In addition, PRS is useful for predicting diseases other than those in the perinatal period.^38, 39^ Given the unchangeability of genomic information, our study suggests that genomic information should be obtained during the relatively healthy childbearing age rather than during middle or old age to avoid poor outcomes in pregnant women due to inadequate prediction of PE. We showed that maternal PRS can improve clinical PE prediction models and suggest its application in other perinatal diseases. In addition, we showed that the utility of paternal genomic information in predicting PE is inconsistent across cohorts; therefore, further studies should aim at identifying populations in which paternal PRS has predictive information for PE.

### Novelty and Relevance

What is New?

- Maternal and paternal genomic information improves clinical prediction models for preeclampsia.

What is Relevant?

- Preeclampsia is a multisystem disorder characterized by de novo hypertension and proteinuria, affecting approximately 3.4% of pregnant women in the USA and 2.7% in Japan.
- Early detection of high-risk pregnancies of preeclampsia can lead to better outcomes for mothers and fetuses. Therefore, developing accurate prediction models is critical in clinical practice.

Clinical/Pathophysiological Implications

- This study provides new insights into the utility of parental genomic information for predicting preeclampsia, which facilitates personalized antenatal care and prevention strategies.

## Data Availability

Individual cohort and genotyping data are available upon request after the approval of the Ethical Committee and the Materials and Information Distribution Review Committee of Tohoku Medical Megabank Organization.

## Acknowledgements

The authors would like to thank all the participants who consented to participate in this study and all the staff at the Tohoku Medical Megabank Organization, Tohoku University, Iwate Tohoku Medical Megabank Organization, and Iwate Medical University. A full list of the members of the Tohoku Medical Megabank Organization is available at https://www.megabank.tohoku.ac.jp/english/a230901/.

## Sources of Funding

This work was supported by the Japan Agency for Medical Research and Development (AMED), Japan (Grant Nos. JP19gk0110039, JP17km0105001, JP21tm0124005, and JP21tm0424601) and JSPS KAKENHI (Grant No. JP21K10438).

## Disclosures

KM is an employee of the Ministry of Education, Culture, Sports, Science and Technology, Japan.

## Non-standard Abbreviations and Acronyms

ACOG: American College of Obstetricians and Gynecologists
BMI: body mass index
BP: blood pressure
C+T: clumping and thresholding
CH: chronic hypertension
CI: confidence intervals
DBP: diastolic blood pressure
DM: diabetes mellitus
GWAS: genome-wide association study
HDP: hypertensive disorders of pregnancy
IVF: in vitro fertilization
JPA NEO: Japonica Array NEO
JPA v2: Japonica Array v2
log MoM: log_10_ transformed multiple of the median
MAP: mean arterial pressure
OR: odds ratio
PC: principal components
PE: preeclampsia
PRS: polygenic risk score
SBP: systolic blood pressure
SD: standard deviation
SLE: systemic lupus erythematosus

## Notes

### Author Declarations

Ethical approval was obtained from the Ethics Committee of the Tohoku Medical Megabank Organization (2023-4-025), and informed consent for research participation was obtained from all participants.

